# Elevated plasma matrix metalloproteinases associate with *Mycobacterium tuberculosis* blood stream infection and mortality in HIV-associated tuberculosis

**DOI:** 10.1101/2023.12.12.23299845

**Authors:** NF Walker, C Schutz, A Ward, D Barr, C Opondo, M Shey, PT Elkington, KA Wilkinson, RJ Wilkinson, G Meintjes

**Author notes:** Corresponding Author:* Dr Naomi F. Walker Department of Clinical Sciences Liverpool School of Tropical Medicine Pembroke Place Liverpool L3 5QA, UK. Authors contributed equally (NF Walker, C Schutz).

## Abstract

Mortality from HIV-associated tuberculosis (HIV-TB) is high, particularly among hospitalised patients. In 433 people living with HIV admitted to hospital with symptoms of TB, we investigated plasma matrix metalloproteinases (MMP) and matrix-derived biomarkers in relation to TB diagnosis, mortality and *Mycobacterium tuberculosis* (*Mtb)* blood stream infection (BSI). Compared to other diagnoses, MMP-8 was elevated in confirmed TB and in *Mtb*-BSI, positively correlating with extracellular matrix breakdown products. Baseline MMP- 3, -7, -8, -10 and procollagen III N-terminal propeptide (PIIINP) associated with *Mtb*-BSI and 12-week mortality. These findings implicate MMP dysregulation in pathophysiology of advanced HIV-TB and support MMP inhibition as a host-directed therapeutic strategy for HIV- TB.

## Background

Tuberculosis (TB) is a leading infectious cause of death worldwide, resulting in an estimated 1.3 million deaths annually (1). The End TB Strategy has highlighted prevention of TB deaths as a major target and aims to reduce TB deaths by 95% between 2015 and 2035 (2). In people living with HIV, TB is the leading cause of death (3). To meet End TB targets, identification of and interventions for those at highest risk of poor outcomes are needed, particularly for use in low-resource settings where the burden of TB falls most heavily. Further understanding of the causes of mortality and pathophysiology of TB disease is required to prevent TB deaths in people living with HIV.

We have previously reported that plasma matrix metalloproteinase (MMP)-1, MMP-8 (neutrophil collagenase) and procollagen N-terminal propeptide (PIIINP), a matrix degradation product (MDP) released during collagen turnover, are elevated in patients with active TB compared to patients without TB, highlighting collagen turnover as a feature of TB disease, in both HIV positive and negative cohorts (4, 5). Plasma PIIINP was significantly higher in HIV positive patients with newly diagnosed active TB compared to HIV negative patients, positively correlated with HIV viral load and was elevated during TB immune reconstitution inflammatory syndrome (IRIS) (4).

Here, we evaluated plasma MMP, PIIINP and extracellular matrix components, collagen type IV alpha 1 chain (Col4⍺1) and hyaluronic acid, in a cohort of hospitalised patients with advanced HIV and TB symptoms, thereby focusing on the population most requiring interventions to reduce mortality. We aimed to evaluate potential as diagnostic biomarkers and hypothesised that disseminated *Mycobacterium tuberculosis* (*Mtb*) in HIV-TB may drive systemic MMP upregulation and consequently tissue damage. We report a novel association of elevated plasma MMP with *Mtb*-blood stream infection (BSI) and mortality in advanced HIV-TB, providing pathophysiological insights.

## Methods

The study was approved by the University of Cape Town Human Research Ethics Committee (HREC ref 057/2013) and London School of Hygiene and Tropical Medicine Research Ethics Committee (ref 11710). Full methods have been reported elsewhere (6). Eligible patients were adults with HIV infection and a CD4 count ≤ 350 cells/μl, admitted to Khayelitsha Hospital, Cape Town, with a probable new diagnosis of TB. Exclusion criteria were pregnancy, TB treatment within one month prior to admission or more than 3 doses of TB treatment prior to enrolment, or unknown HIV status (declined testing). Participants provided written informed consent. Eligible patients with decreased capacity to consent were enrolled and followed up daily to obtain consent according to the approved protocol. Participants were investigated by TB culture (sputum, blood), Xpert (sputum, urine) and urine lipoarabinomannan (LAM, Alere Determine TB LAM assay) prior to initiation of TB treatment. All results were made available to the clinical team and participants remained in routine clinical care. Vital status was determined at 12 weeks.

Participants were classified retrospectively as microbiologically **confirmed TB** if *Mtb* was identified in clinical samples, **probable TB** if TB was likely and treatment for TB was given following WHO guidelines but no microbiological confirmation was obtained, **no TB**, if TB was excluded on clinical and microbiological grounds, or **LAM TB** if criteria for no TB was met but urine LAM was positive >=2 by two independent readers (6).

Inclusion in this analysis required the availability of EDTA plasma samples at enrolment. MMP -1, -3, -7, -8, -9 and -10 were quantified by Luminex array (Bio-Rad Bio-Plex 200 system; assay R&D Systems, United Kingdom). PIIINP, Col4⍺1 and hyaluronic acid were quantified by enzyme-linked immunosorbent assays (Cloud Clone Corp, China) performed as per manufacturers’ instructions. Hyaluronic acid and Col4⍺1 measurement was limited to a subset of 73 randomly selected participants. Statistical analysis was performed in Prism 9 and R Studio (2023.03.1). Comparisons of analytes between groups was Mann-Whitney U analysis unless otherwise stated. Statistical significance was inferred by a p value <0.05. A Bonferroni correction was performed where more than two groups were compared for one analyte.

The relationship between MMP or PIIINP concentrations and *Mtb*-BSI or mortality was assessed using a mixed effects model, including a random effect on intercept for plate. Hierarchical clustering analysis was performed by Ward’s method based on Euclidean distance, to assess the association between analytes, including neutrophil count, neutrophil percentage and procalcitonin, but excluding Col4⍺1 and HA, as they had only been measured in a subset of patients. This was on scaled data (mean subtracted and divided by standard deviation), excluding extreme outliers (observations with a value greater than four median absolute deviations from the variable median applied after transformation). Pearson correlation to assess the association between MMP and extracellular matrix breakdown product concentrations was performed on log-transformed values excluding extreme outliers as above.

## Results

### Participant demographics, diagnoses and outcomes

Plasma samples were available for 437 participants. The median CD4 count was 62 cells/mm^3^ (IQR 22.5-133). Participant demographics are reported in Supplementary Table S1. TB diagnosis was confirmed in 313 (71.6%) and clinical TB in 48 (11.0%). Only 4 participants met the criteria for LAM TB, so these were excluded from laboratory analyses below. In 72 participants who had TB excluded (no TB), community acquired pneumonia (n=34, 47.2%) was the most frequent alternative diagnosis, followed by other blood pathogen (n=8, 11.1%), *Pneumocystis jirovecii* pneumonia (n=6, 8.33%) and cryptococcal disease (n=6, 8.33%).

Vital status at 12 weeks was known for 431/437 (98.6%) participants. Death occurred in 83/437 (19.0%) participants at a median of 16.0 (IQR 3.0-43.0) days. Mortality at 12 weeks in confirmed TB was 19.8% (62/313). *Mtb*-BSI was present in 133/313 (42.5%) participants with confirmed TB and was associated with increased mortality at 12 weeks: 37/133 (27.8%) participants with *Mtb*-BSI died compared to 23/173 (13.3%) without (p = 0.002 by Fisher’s Exact Test).

### Plasma MMP-8 is elevated in patients hospitalised with HIV-associated TB

We first examined plasma MMP and extracellular matrix breakdown product concentrations in confirmed TB in comparison to clinical TB or no TB. Plasma MMP-8 was significantly elevated in confirmed TB compared to no TB (median 23712 pg/ml, IQR 7688-47571 vs median 10602, IQR 2019-32205; p=0.003; Figure 1A and Supplementary Table S2), as was plasma Col4⍺1 (Figure 1B). Plasma MMP-3 and -10 were lower in participants with confirmed TB compared to no TB and clinical TB, whilst MMP-1, -7 and -9, PIIINP and hyaluronic acid did not differ between groups (Supplementary Figure S1).

**Figure 1.**
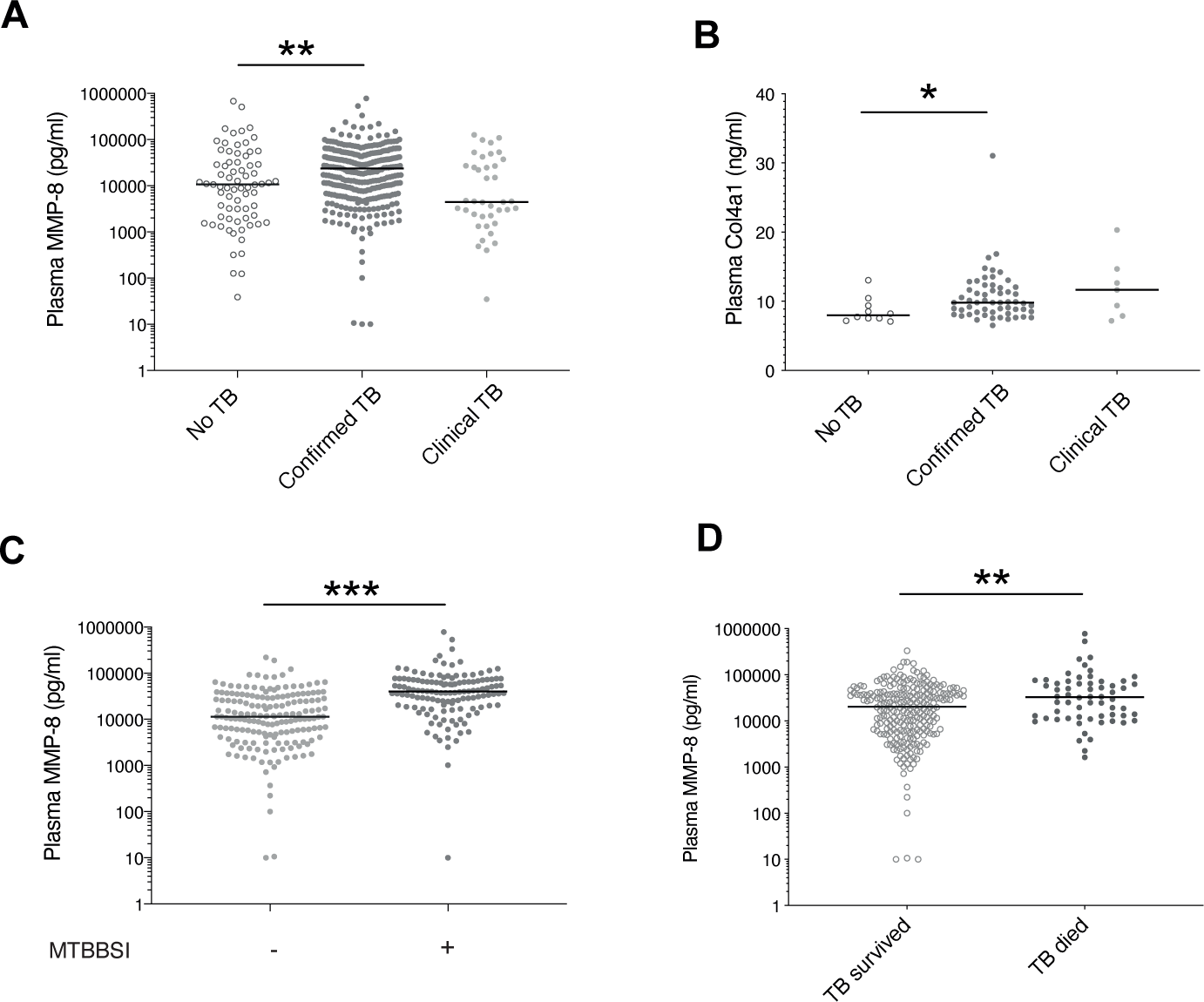
Elevated matrix metalloproteinase-8 in HIV-TB, *Mtb*-blood stream infection and TB mortality Plasma matrix metalloproteinase (MMP)-8 (A) and Col4⍺1 (B) were elevated in hospitalised participants with HIV infection and microbiologically-confirmed TB, compared to hospitalised HIV positive participants with symptoms due to other diagnoses (no TB). Differences between patients with confirmed TB and TB diagnosed clinically were not statistically significant. Plasma MMP-8 was increased in participants with confirmed TB and *Mycobacterium tuberculosis* blood stream infection (MTBBSI) compared to those without (C) and in those who had died compared to those who had survived at 12 weeks (D). Col4⍺1 was measured in a subset of 73 participants. *p<0.05, **p<0.01, ***p<0.001.

### Plasma MMP-8 is associated with Mtb blood stream infection and mortality in HIV-associated TB

MMP are primarily transcriptionally regulated however, MMP-8 may be stored in neutrophil granules. Exploring the hypothesis that disseminated *Mtb* drives MMP-8 upregulation and release from neutrophils and that this associates with poor outcomes in HIV-TB, we next examined MMP-8 concentration in the presence or absence of *Mtb*-BSI and by vital status at 12 weeks. We found that amongst participants with confirmed TB, those with *Mtb*-BSI had elevated plasma MMP-8 (Figure 1C) compared to those without (median 40003 pg/ml, IQR 20006-70583 vs median 11451, IQR 4697-30789; p<0.001). Plasma MMP-8 was elevated in participants with confirmed TB who died compared to those who survived (median 32811 pg/ml IQR 12060-66934 vs median 20201, IQR 6050-40561, p=0.002, Figure 1D). Col4⍺1 and hyaluronic acid concentrations did not differ between those who died and those who survived (data not shown), although Col4⍺1 was elevated in patients with confirmed TB with *Mtb-*BSI compared to those without (median 11.2, IQR 7.76-9.92 vs 8.75ng/ml IQR 9.76-13.5, p<0.001). Plasma MMP-8 positively correlated with Col4⍺1 (Pearson r = 0.535, p<0.001) and PIIINP (Pearson r = 0.404, p<0.001) but not with hyaluronic acid concentrations.

### Multiple MMP and PIIINP associate with HIV-TB severity

We proceeded to examine the association of other plasma MMP and PIIINP with probability of *Mtb*-BSI and mortality in confirmed TB. We found a positive association between plasma MMP-3, -7, -8, -10 and PIIINP for Mtb-BSI and mortality (Figure 2A and 2B). We performed hierarchical clustering analysis including neutrophil count, neutrophil percentage, and procalcitonin as biomarkers of acute inflammation in addition to MMP concentrations and PIIINP (Supplementary Figure S1). We found that MMP-8 most closely clustered with PIIINP and procalcitonin. Procalcitonin was positively associated with *Mtb*-BSI and mortality, but neutrophil count was not (Figure 2C).

**Figure 2.**
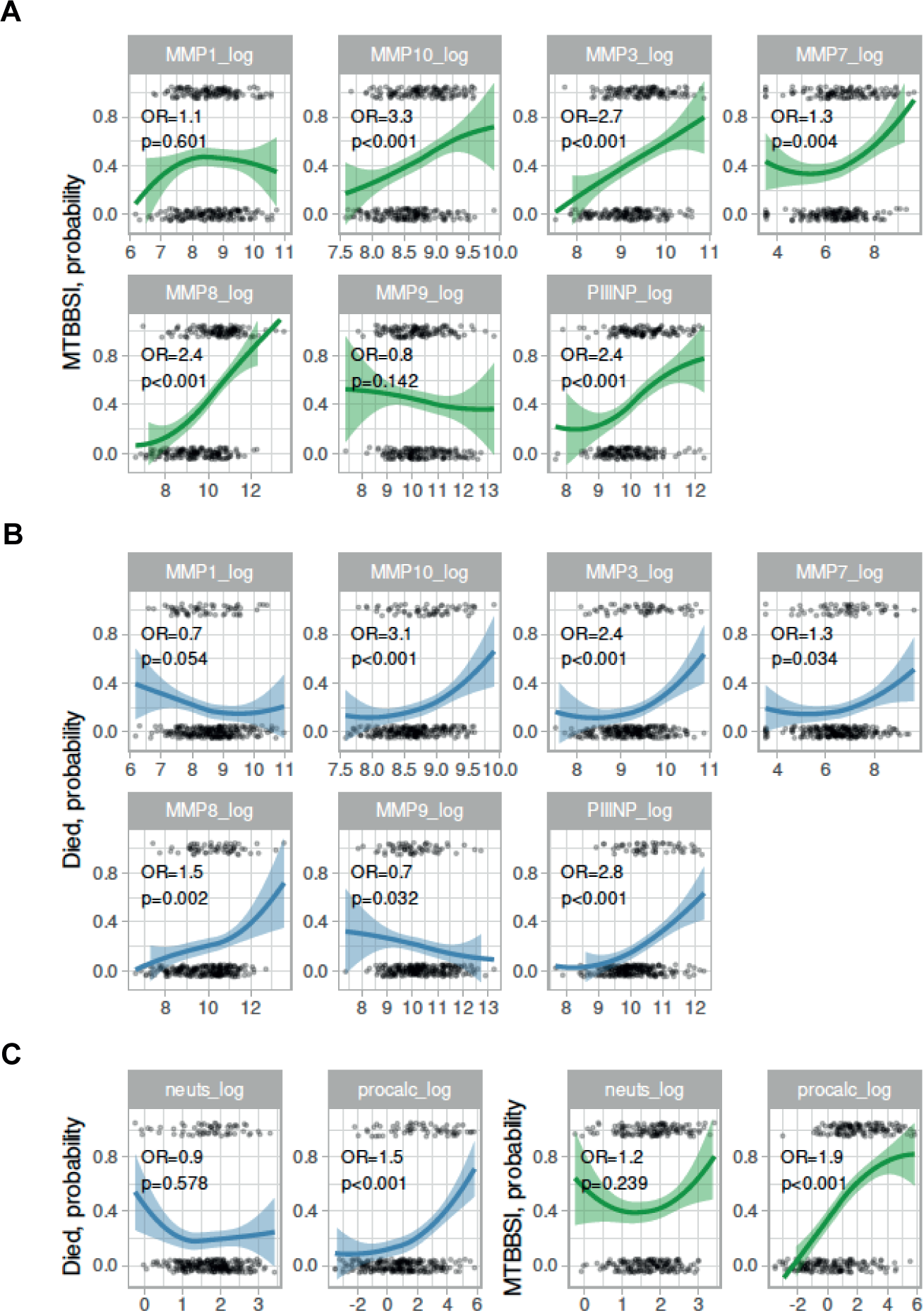
Plasma matrix metalloproteinase (MMP) and procollagen III N-terminal propeptide (PIIINP) associate with *Mycobacterium tuberculosis* blood stream infection and mortality in HIV-associated TB. A and B: Analyte (log concentration, x-axes) associations (y-axes) with Mycobacterium tuberculosis blood stream infection (MTBBSI) and mortality. Odds ratios (OR) and p-values are from logistic regression models, with adjustment for random effects by plate. C: Log neutrophil and procalcitonin (x-axes) associations (y-axes) with MTBBSI and mortality. All panels: Loess fit to data is shown (coloured lines with shaded 95% confidence intervals).

## Discussion

Patients with advanced HIV-1 hospitalised with symptoms of TB are at high risk of mortality. In this study, we found that baseline plasma MMP-8 and Col4⍺1 were increased in hospitalised patients with HIV infection who were confirmed to have TB compared to those who eventually received an alternative diagnosis. Contrary to our previous finding in outpatients, in this hospitalised cohort, plasma PIIINP was not elevated in patients with TB compared to symptomatic patients with alternative diagnoses.

Elevated plasma MMP-8, together with MMP-3, -7, -10 and PIIINP associated with *Mtb*-BSI and mortality at 12 weeks, suggesting that MMP upregulation and matrix turnover are features of TB disease severity. This is in keeping with our recently reported finding that elevated plasma MMP-8 concentrations at the end of TB treatment are associated with persistent *Mtb* culture positivity (7). In the clustering analysis, MMP-8 was most closely associated with procalcitonin (an acute phase reactant) and the matrix degradation product, PIIINP (released during Type III collagen turnover), suggesting that MMP-8 activity, collagen turnover and acute inflammation are closely related processes in HIV-TB (5, 8). Hyaluronic acid, a polysaccharide component of the extracellular matrix which is released during extracellular matrix turnover has previously been found to predict AIDS events or death in HIV positive patients commencing antiretroviral therapy (9). Plasma hyaluronic acid was not associated with mortality in this study.

Whilst we have demonstrated an association between plasma MMP and mortality in HIV-TB, our study does not prove a causal relationship. Elevated MMP may be a function of *Mtb* bacterial load or Mtb dissemination, which is itself associated with mortality. *In vitro*, virulent *Mtb* is able to induce neutrophil-derived MMP-8 secretion, via an NF-kB-dependent mechanism, so it is possible that blood stream *Mtb* directly stimulates neutrophil MMP-8 release (10). However, elevated MMP-8 may also occur via upregulation of pro-inflammatory cellular networks and the finding that MMP-8 associated more closely with procalcitonin than neutrophil count or neutrophil percentage consistent with networks upregulating MMP-8 (11). Increased MMP-8 in TB was positively associated with PIIINP, a matrix degradation product released during type III collagen turnover and Col4⍺1, a component of type IV collagen. MMP inhibition with doxycycline has been shown to be safe and effective for TB in HIV negative patients (12). Our results support the case for clinical trials of MMP inhibition with doxycycline as a host-directed therapy for HIV-associated TB.

A number of prior studies have examined MMP-8 concentrations in patients with TB, predominantly in those who are HIV negative (11). Our finding of elevated plasma MMP-8 in participants with confirmed TB compared to those symptomatic with other illnesses is consistent with previous reports in outpatients, although this is the first report in people who are HIV positive and hospitalised (4, 5, 13, 14). MMP-8 is emerging as a biomarker that may help identify patients with active TB, in some settings if used in combination with other screening tools. Here, we also report elevated plasma Col4⍺1 in HIV-TB and elevated Col4⍺1 was also associated with *Mtb*-BSI. Col4⍺1 is a subunit of Type IV collagen, a key component of basement membranes that has not previously been studied in human TB, to our knowledge.

A strength of this study is the combination of rigorous clinical and mycobacterial analyses that was employed to determine TB status. A limitation pertains to the use of a single blood culture to determine the presence or absence of *Mtb*-BSI. It is likely that sequential blood cultures would have a higher diagnostic accuracy for *Mtb*-BSI (15).

In summary, in hospitalised people living with HIV, with a clinical syndrome compatible with TB, we have reported elevated MMP-8 in confirmed TB, likely due to *Mtb*-driven tissue damage, compared to other diagnoses. Furthermore, plasma MMP-3, -7, -8, -10 and PIIINP associated with *Mtb*-BSI and mortality at 12 weeks, implicating MMP dysregulation in HIV-TB morbidity. MMP inhibition is a potential therapeutic target for this patient group, who are at urgent need of improved therapeutic strategies.

## Data Availability

All laboratory data in the present study are available upon reasonable request to the authors

## Acknowledgments

We thank the Western Cape Provincial Government and staff at Khayelitsha Hospital for their support of the study. NFW was supported by an NIHR Academic Clinical Lectureship and funding from Academy of Medical Sciences UK, MRC UK, British Heart Foundation, Arthritis Research UK, Royal College of Physicians and Diabetes UK (Starter Grant for Clinical Lecturers) and British Infection Association (Project Grant). MS is supported by Wellcome (211360/Z/18/Z) and the National Research Foundation of South Africa (NRF, #UID127558). PTE was supported by MRC MR/P023754/1 and MR/W025728/1. RJW is supported by the Francis Crick Institute which is funded by Wellcome (CC2112), Cancer Research UK (CC2112) and UKRI (CC2112). RJW also receives support from Wellcome (203135), EDCTP (SRIA 2015- 1065) and NIH (U01AI115940). CS was funded by the South African Medical Research Council under the National Health Scholars Programme. GM was supported by Wellcome (098316, 214321/Z/18/Z, and 203135/Z/16/Z), and the South African Research Chairs Initiative of the Department of Science and Technology and National Research Foundation (NRF) of South Africa (Grant No 64787). The funders had no role in the study design, data collection, data analysis, data interpretation, or writing of this report. The opinions, findings and conclusions expressed in this manuscript reflect those of the authors alone.

This research was funded in whole, or in part, by Wellcome [098316, 214321/Z/18/Z, 203135/Z/16/Z]. For the purpose of open access, the author has applied a CC-BY public copyright licence to any Author Accepted Manuscript version arising from this submission.

## Author contributions

GM, CS, AW, DB conceived the clinical study and recruited the clinical cohort. NFW, GM, PE, DB conceived the laboratory study. NFW, KAW, MS, DB conducted laboratory analyses. NFW, CS, CO, DB performed data analysis. NFW wrote the first draft of the manuscript. All authors contributed to the manuscript and approved the final submitted report.

## List of Supplementary Tables and Figure

Supplementary Table S1. Demographic and Clinical Features of Study Participants

Supplementary Table S2. Matrix metalloproteinase and extracellular matrix breakdown product concentrations

Supplementary Figure S1 Plasma matrix metalloproteinase-8 associates with collagen turnover and acute inflammation

