## Supplemental Tables and Figure for "Elevated plasma matrix metalloproteinases associate with *Mycobacterium tuberculosis* blood stream infection and mortality in HIV-associated tuberculosis"

Supplementary Tables and Figure

**Supplementary Table S1 Demographic and Clinical Features of Study Participants**

| | Non-TB | Confirmed TB | Clinical TB | LAM TB | All participants |
|---|---|---|---|---|---|
| Frequency, n (%) | 72 (16.5) | 313 (71.6) | 48 (11.0) | 4 (0.9) | 437 (100) |
| Median age (years), IQR | 39.9 (30.9-49.3) | 35.7 (30.5-43.0) | 38.0 (31.6-42.5) | 36.8 (34.3-37.9) | 36.1 (30.8-43.9) |
| Female, n (%) | 40 (55.6) | 159 (50.8) | 29 (60.4) | 2 (50.0) | 230 (52.6) |
| Median CD4 count (cells/mm$^3$), IQR | 89.5 (29.0-224) | 56 (18.0-112) | 93.0 (53.8-182) | 88.0 (26.8-137) | 62.0 (22.5-133) |
| On ART, n (%) | 28 (38.9) | 95 (30.4) | 19 (39.6) | 0 (0) | 142 (32.5) |
| Mortality at 12 weeks, n (%) | 11 (15.3) | 62 (19.8) | 9 (18.8) | 1 (25.0) | 83 (19.0) |

Abbreviations: ART = antiretroviral therapy; IQR = interquartile range; LAM = lipoarabinomannan.

**Supplementary Table S2 Matrix metalloproteinase and extracellular matrix breakdown product concentrations**

| | No TB | | Confirmed TB | | Clinical TB | | p values$ | |
|---|---|---|---|---|---|---|---|---|
| Plasma | Median | IQR | Median | IQR | Median | IQR | No TB vs Confirmed TB | No TB vs Clinical TB |
| MMP-1 (pg/ml) | 4477 | 1988, 7640 | 5316 | 2686, 9756 | 5163 | 2671, 9799 | 0.112 | 0.162 |
| MMP-3 (pg/ml) | 12840 | 8180, 22951 | 10716 | 6934, 15935 | 8535 | 5528, 14080 | **0.021** | **0.006** |
| MMP-7 (pg/ml) | 1071 | 455, 1967 | 712 | 330, 1406 | 525 | 174, 1589 | 0.046 | 0.088 |
| MMP-8 (pg/ml) | 10602 | 2019, 32205 | 23712 | 7688, 47571 | 4430 | 2156, 27099 | **0.003** | 0.324 |
| MMP-9 (pg/ml) | 26512 | 11079, 45181 | 26574 | 14400, 58120 | 23853 | 12909, 50802 | 0.220 | 0.667 |
| MMP-10 (pg/ml) | 7809 | 4990, 11089 | 5807 | 4147, 8009 | 5310 | 3798, 7897 | **<0.001** | **0.001** |
| PIIINP (pg/ml) | 21380 | 14323, 41866 | 22712 | 13978, 40724 | 19308 | 12082, 28705 | 0.941 | 0.193 |
| HA (ng/ml) | 8.26 | 5.24, 12.9 | 8.287 | 5.815, 10.88 | 8.815 | 5.29, 15.5 | 0.640 | 0.872 |
| Col4α1 (ng/ml) | 7.98 | 7.45, 9.65 | 9.81 | 8.42, 12.0 | 11.7 | 7.87, 14.7 | 0.017 | 0.103 |

Confirmed TB*: Mycobacterium tuberculosis* identified in clinical samples; Clinical TB*:* TB was likely and the patient was treated for TB but no microbiological confirmation was obtained; No TB: TB was excluded on clinical and microbiological grounds. Abbreviations: Col4α1 = collagen IV alpha 1; HA = Hyaluronic Acid (HA); IQR = interquartile range (IQR); matrix metalloproteinase (MMP); PIIINP = procollagen N-terminal propeptide. HA and Col4α1 measurement were on a random subgroup of 73 participants.

$A Bonferroni correction for multiple comparisons indicated that a p value of <0.025 was equivalent to a significance threshold of <0.05. Significant p values are shown in bold.

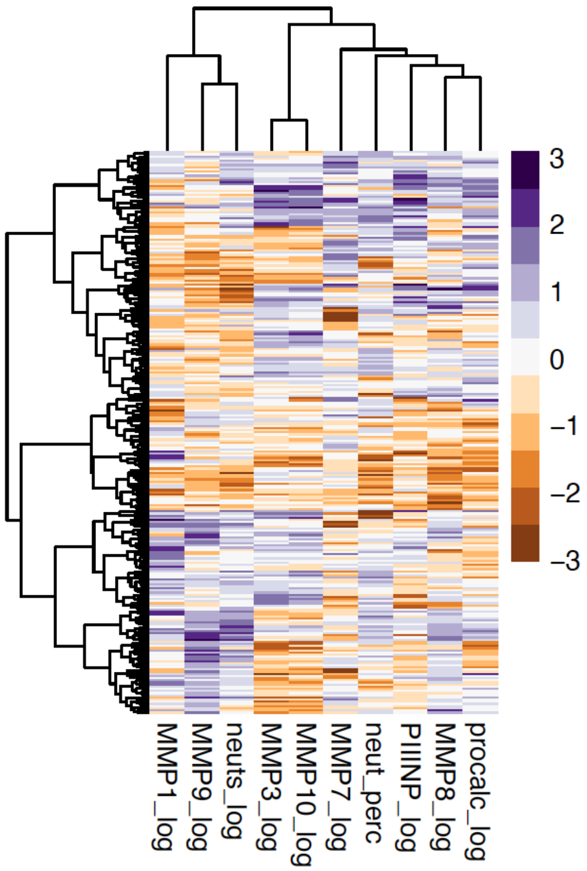


**Supplementary Figure S1 Plasma matrix metalloproteinase-8 associates with collagen turnover and acute inflammation**

Hierarchical clustering analysis demonstrates that plasma matrix metalloproteinase-8 (MMP-8) most closely associated with procalcitonin (procalc) concentrations. Plasma MMP-3 and MMP-10 clustered together, whilst MMP-9 mostly closely clustered with neutrophil count (neuts). The analysis was on scaled data, excluding extreme outliers, which were less than ten data points for any one analyte. Neuts_perc = neutrophil percentage; PIIINP = procollagen III N-terminal propeptide.
